# Can serum biomarkers predict the outcome of systemic therapy for atopic dermatitis?

**DOI:** 10.1101/2020.12.02.20242404

**Authors:** Guillem Hurault, Evelien Roekevisch, Mandy E. Schram, Krisztina Szegedi, Sanja Kezic, Maritza A. Middelkamp-Hup, Phyllis I. Spuls, Reiko J. Tanaka

**Affiliations:** Department of Bioengineering, Imperial College London; Amsterdam UMC, location AMC, University of Amsterdam, Department of Dermatology, Amsterdam Public health, Infection and Immunity

## Abstract

**Background:** Atopic dermatitis (AD or eczema) is a most common chronic skin disease. Designing personalised treatment strategies for AD based on patient stratification, rather than the “one-size-fits-all” treatments, is of high clinical relevance. It has been hypothesised that the measurement of biomarkers could help predict therapeutic response for individual patients.

**Objective:** We aim to assess whether biomarkers can predict the outcome of systemic therapy.

**Methods:** We developed a statistical machine learning predictive model using the data of an already published longitudinal study of 42 patients who received systemic therapy. The data contained 26 serum cytokines measured before the therapy. The model described the dynamics of the latent disease severity and measurement errors to predict AD severity scores (EASI, (o)SCORAD and POEM) two-weeks ahead. We conducted feature selection to identify the most important biomarkers for predicting the AD severity scores.

**Results:** We validated our model and confirmed that it outperformed standard time-series forecasting models. Adding biomarkers did not improve predictive performance. Our estimates of the minimum detectable change for the AD severity scores were larger than already published estimates of the minimal clinically important difference.

**Conclusions:** Biomarkers had a negligible and non-significant effect for predicting the future AD severity scores and the outcome of the systemic therapy. Instead, a historical record of severity scores provides rich and insightful dynamical information required for prediction of therapeutic responses.

## INTRODUCTION

Atopic dermatitis (AD, also called eczema) is one of the complex diseases with a considerable variation in the clinical phenotype and responses to treatments among patients [1]. Current treatments aim to manage chronic AD symptoms by preventing exacerbations, mainly using emollients, topical corticosteroids, calcineurin inhibitors, systemic therapies and biologics. Given the heterogeneity in responses to different treatments and AD phenotypes, it is of high clinical relevance to stratify patients and to design personalised treatment strategies for AD rather than using “one-size-fits-all” treatments [2] [3].

The identification of biomarkers of AD has been considered to be a critical step toward precision medicine [3]. Thymus and activation-regulated chemokine (TARC) was suggested to be the single best biomarker to assess disease severity [4], panels of biomarkers were proposed as “objective” substitutes for the EASI [5] and SCORAD [6] severity scores, and a discovery of AD endotypes was attempted by clustering of biomarker measurements [7]. Furthermore, the presence of FLG mutations [9] [10] and a high level of serum IgE [11] were found to be associated with poor treatment outcome for AD. However, biomarkers to *predict* therapeutic responses has been less studied for AD [8]. Generating predictions of *future* AD severity, beyond quantifying associations, is crucial to make patient stratification clinically relevant for personalised medicine. Ideally, the effects of multiple biomarkers should be investigated in a multivariable regression and predict therapeutic responses at more than one timepoint.

We recently developed a statistical machine learning model to predict daily AD severity scores at an individual patient level [12]. While our model investigated the effects of age, ethnicity and treatment usage on predictive performance, we could not investigate the predictive power of biomarkers (other than the presence of filaggrin mutation) due to the unavailability of such data. To examine whether biomarkers can predict future AD severity scores, in this study, we developed a Bayesian state-space model to predict the evolution of AD severity scores using the longitudinal data of AD patients under a systemic therapy collected in an already published study [13] [14]. We applied feature selection to identify potential biomarkers for patient stratification.

## METHODS

### Data

We used the data from an already published longitudinal clinical study [13] [14] where 42 adult AD patients received systemic therapy with either azathioprine (AZA) or methotrexate (MTX) for over 24 weeks. The concentrations of 26 serum biomarkers were measured for each of 42 patients before the start of the treatment (week 0). The values were log-transformed and standardised to have a mean 0 and a variance 1. Three out of 1092 (= 26 ⨯ 42) measurements of the biomarkers were missing and imputed by the population mean value of the corresponding biomarker.

The AD severity for each patient was assessed by SCORAD [15], oSCORAD (the objective component of SCORAD), EASI [16] and POEM [17] at weeks 0, 2, 4, 8, 12 and 24 from the start of the therapy. Our model assumed a constant interval of two weeks from week 0 to week 24 and treated the absence of the AD severity measurement at weeks 6, 10, 14, 16, 18, 20 and 22 as missing values. It resulted in 56.2% missing values for EASI. Patients were genotyped for mutations in the gene coding filaggrin. The missing filaggrin mutation status for six patients was imputed by “no mutation”. The patients’ age was standardised to have a mean 0 and a variance 1.

### Model overview

We developed a Bayesian state-space model to make probabilistic predictions of future AD severity scores (either EASI, SCORAD, oSCORAD or POEM). The Bayesian model describes uncertainties in parameters and severity scores as probability distributions. We assume that the observed severity score is an imperfect measurement of the true latent (unobserved) score and model how the latent score changes over time (Fig. 1). Modelling the measurement errors also allows us to estimate the minimum detectable change (MDC), that is “the smallest change that can be considered above the measurement error with a given level of confidence” [18].

**Figure 1:**
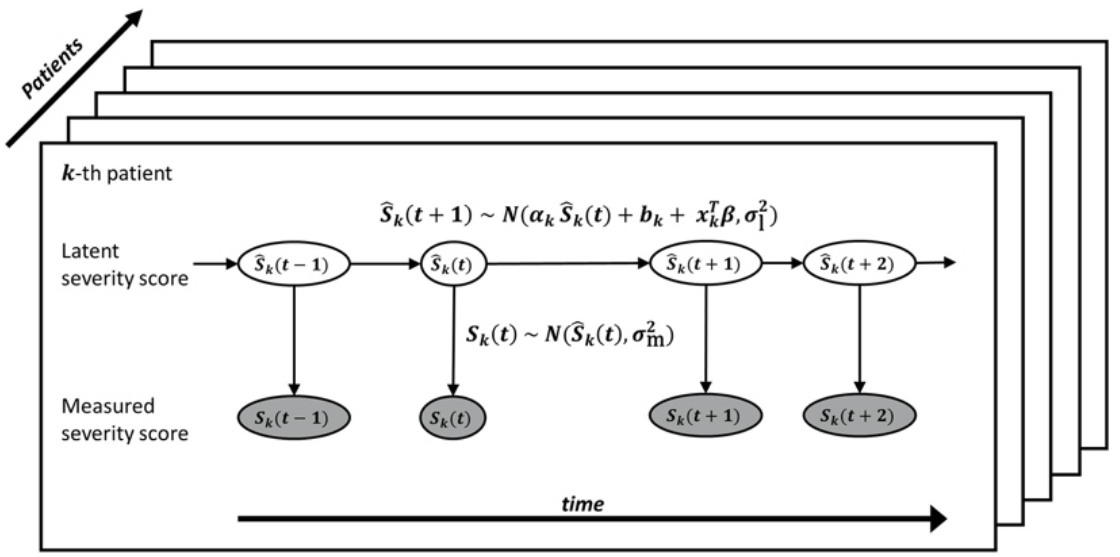
A schematic diagram of the Bayesian state-space model for probabilistic predictions of AD severity scores. Grey and white ovals represent measured and latent (unobserved) scores, respectively.

For the *k*-th patient at time *t*, we assume that the measurement of a score, *S*_*k*_(*t*),is generated from a truncated Gaussian distribution, 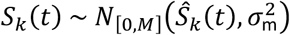,centred around the latent score, *Ŝ*_*k*_(*t*). The distribution is truncated between 0 and the maximum value, *M*,of the severity score (72 for EASI, 83 for oSCORAD, 103 for SCORAD and 28 for POEM). The standard deviation of the measurement process, *σ*_m_, quantifies the measurement error and the minimum detectable change for the default 95% confidence level is determined by *MDC*=1.96*σ*_m_.

The latent dynamics of *Ŝ*_*k*_(*t*) was modelled by a mixed effect autoregressive model, 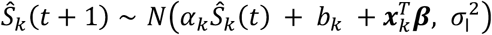, where *α*_*k*_ is the autocorrelation parameter, *b*_*k*_ is the intercept, ***x***_*k*_ is a covariates vector (including biomarkers) with their coefficients, ***β***, and *σ*_l_ is the standard deviation of the latent dynamics. We performed feature selection on the covariates *x*_*k*_ by assuming a regularised horseshoe prior for ***β*** [19]. The horseshoe prior shrinks small coefficients toward 0 while allowing strong signals to remain large, thus limiting overshrinkage unlike *L*_1_ or *L*_2_ regularisations [20]. We assumed a hierarchical prior for *α*_*k*_ and *b*_*k*_ and weakly informative priors for the other parameters (detailed in Supplementary A).

Model inference was performed using the Hamiltonian Monte-Carlo algorithm in the probabilistic programming language Stan [21] with four chains and 2000 iterations per chain including 50% burn-in. Prior predictive checks and fake data checks were conducted. Convergence and sampling were monitored by looking at trace plots, checking the Gelman-Rubin convergence diagnostic 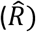 [22], and computing effective sample sizes (*N*_eff_).

### Model validation

The predictive performance of our model was assessed by *K*-fold cross-validation (*K* = 7,stratified by patients) in a forward chaining setting (Fig. S1), to reflect how the model would be used in a clinical setting. For each fold, the model was pre-trained with (*K* − 1) subsets of patients and validated on the remaining subset of patients in a forward chaining setting, in which the model was trained with the first timepoint and tested on the remaining timepoints, then the model was trained with the first two timepoints and tested on the remaining timepoints, etc.

The probabilistic predictions of AD severity scores were evaluated by a logarithmic scoring rule, the log predictive density (lpd). We compared the lpd of our model to that of four reference models, a uniform forecast model (Uniform), a random walk model (RW), an autoregressive model (AR) and a mixed effect autoregressive model (MixedAR). Details of the reference models are described in Supplementary B.

## RESULTS

### Model fit and validation

The Bayesian state-space model that predicts EASI without covariates was fitted to the data successfully. We found no evidence of an absence of convergence. Population-level parameters were estimated with good precision with posterior distributions narrower than their prior distributions (Table S1). We confirmed that the patient-dependent parameters, *α*_*k*_ and *b*_*k*_,vary between patients, within the range of [0.37, 0.99] for the expected autocorrelation (*α*_*k*_) and [0.03, 2.3] for the expected intercept (*b*_*k*_). The measurement process is responsible for 94.7% (90% credible interval 87.3-99.1%) of the total variance for prediction. The posterior mean of the minimum detectable change, *MDC*, is 8.6 (90% credible interval 7.6-9.6). The posterior predictive distribution of EASI trajectories demonstrated that the model could capture different patterns, despite the absence of several measurements (Fig. 2).

**Figure 2:**
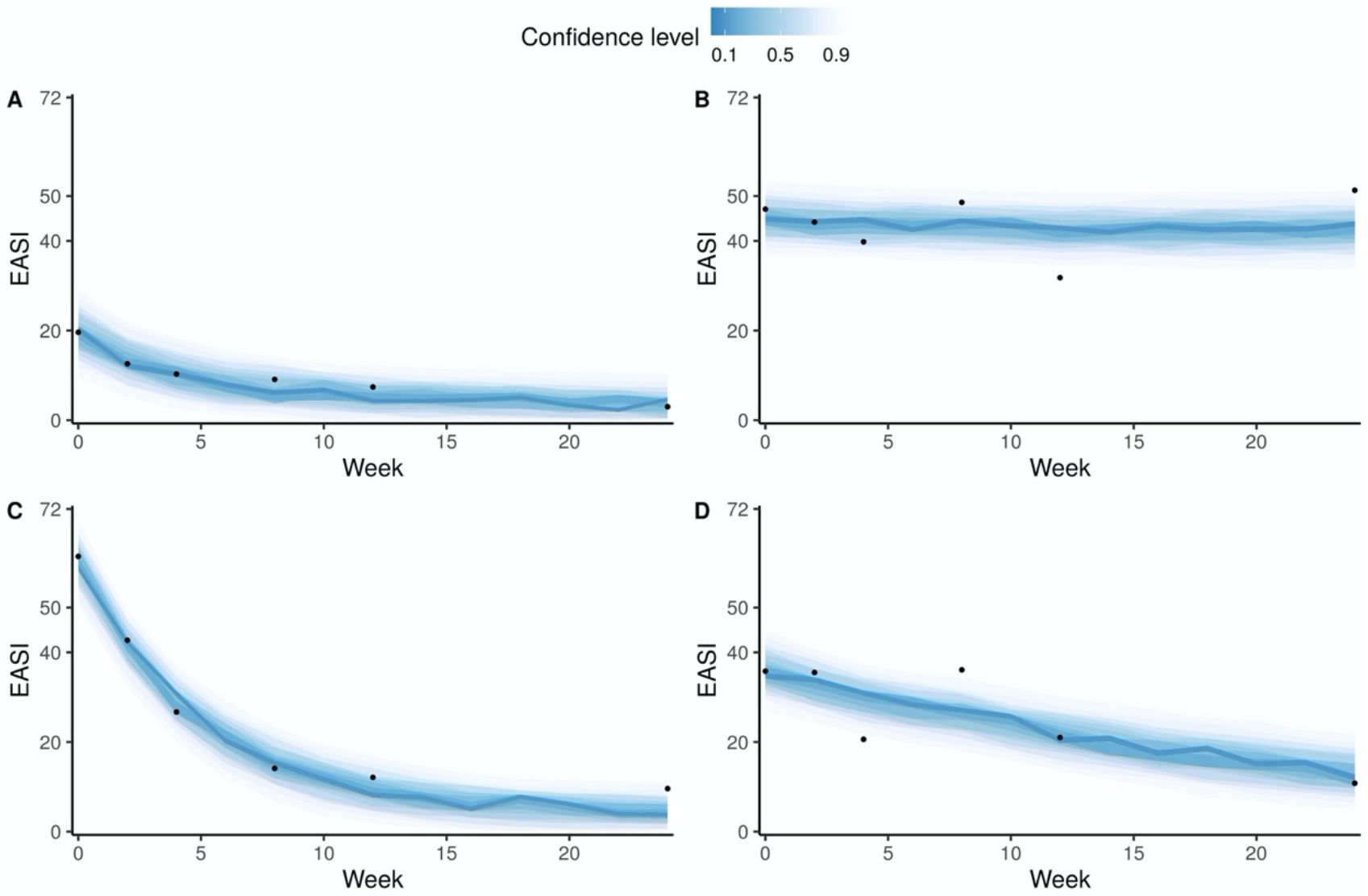
The posterior predictive distribution of four representative patients (A-D) by our model. Dots indicate the measured EASI scores, and the coloured ribbons represent stacked credible intervals. Lighter and darker ribbons correspond to wider and narrower highest density credible intervals, respectively. **A**: Slow recovery from a moderate EASI. **B**: Persistent severe EASI. **C**: Rapid recovery from a severe EASI. **D**: Slow recovery from a severe EASI.

Learning curves for two-weeks ahead predictions (Figs 3A and S2) demonstrated that the predictive performance improved as more data came in and that our state-space model (SSM) outperformed the reference models, thus supporting our model structure. The root mean squared error of the mean prediction for EASI at the next clinical visit (e.g. from week 0 to 2, 2 to 4, 4 to 8, etc.) was 6.3 ± 0.62 for our model and 9.9 ± 0.43 for the random walk model. Counterintuitively, the performance of our model and the mixed autoregressive model tend to improve as the prediction horizon increased (Fig. 3B), possibly because most patients tend to recover before the end of the study.

**Figure 3:**
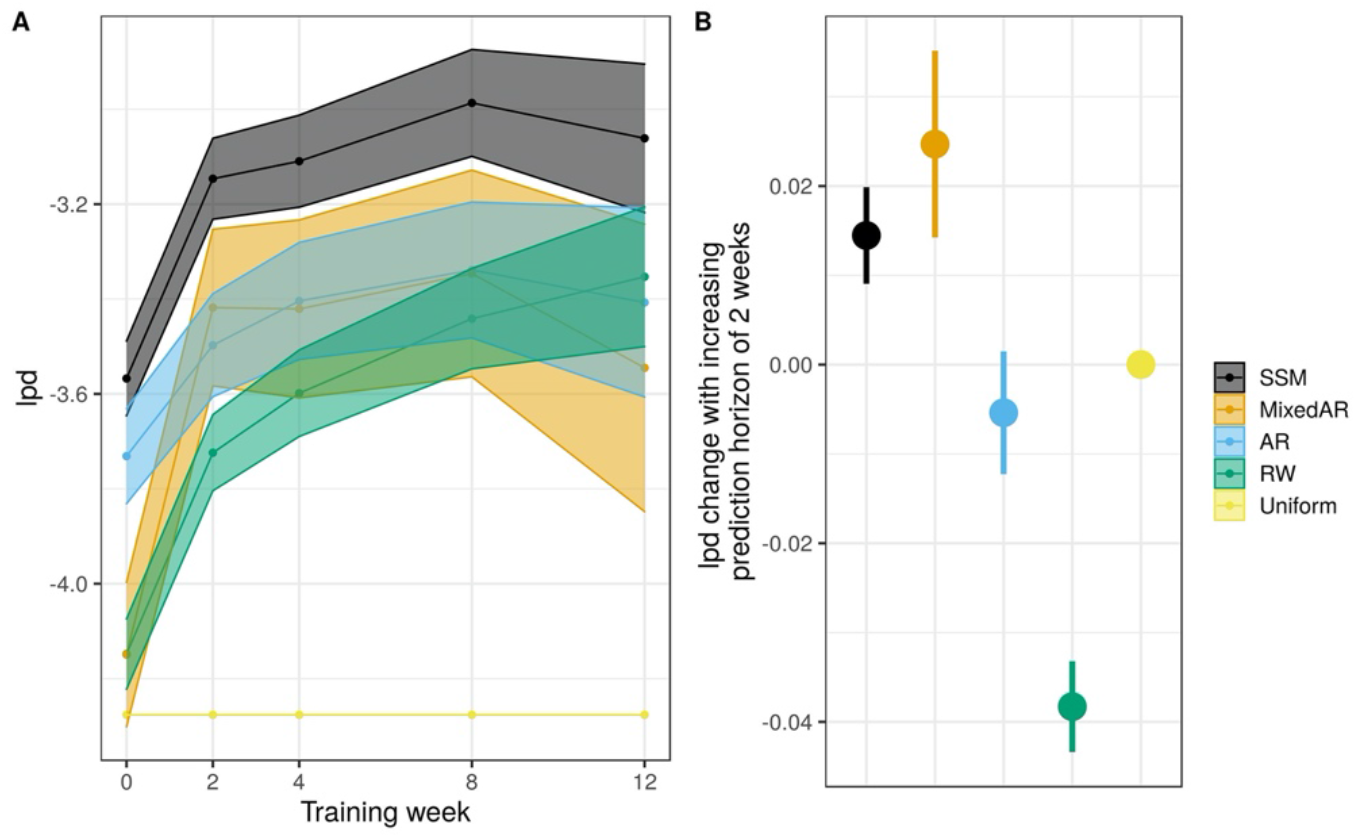
Performance of our model (SSM) and reference models (MixedAR, AR, RW and Uniform) to predict EASI. The performance was evaluated by lpd (higher the better). **A:** Learning curves (mean ± SE) for two-weeks ahead prediction after adjusting for different prediction horizons. **B:** Change in lpd as the prediction horizon is increased by two weeks.

Similar results, with lower performance relative to the reference models, were obtained for the model predicting oSCORAD, SCORAD and POEM (Fig. S3). The posterior means (and 90% credible intervals) of the *MDC* were 9.1 (7.4-10.7) for oSCORAD, 11.4 (9.1-13.5) for SCORAD and 7.7 (6.7-8.9) for POEM.

### Effect of biomarkers on the model’s predictions

Our model that predicts EASI with covariates was also fitted successfully. The covariates included the 26 serum biomarkers measured at week 0, the presence of filaggrin mutation, the systemic therapy applied (AZA or MTX), sex and age. None of the covariates had a practically significant effect on the model’s prediction, as indicated by a small magnitude of the posterior mean and 90% credible intervals on both sides of 0 (Fig. 4A), and a resulting small and not practically significant contribution of the covariate, 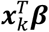,to the EASI prediction (Fig. 4B). The predictive performance of the model with covariates was similar to that of the model without covariates. Similarly, we found no practically significant covariates for the predictive models of oSCORAD, SCORAD and POEM.

**Figure 4:**
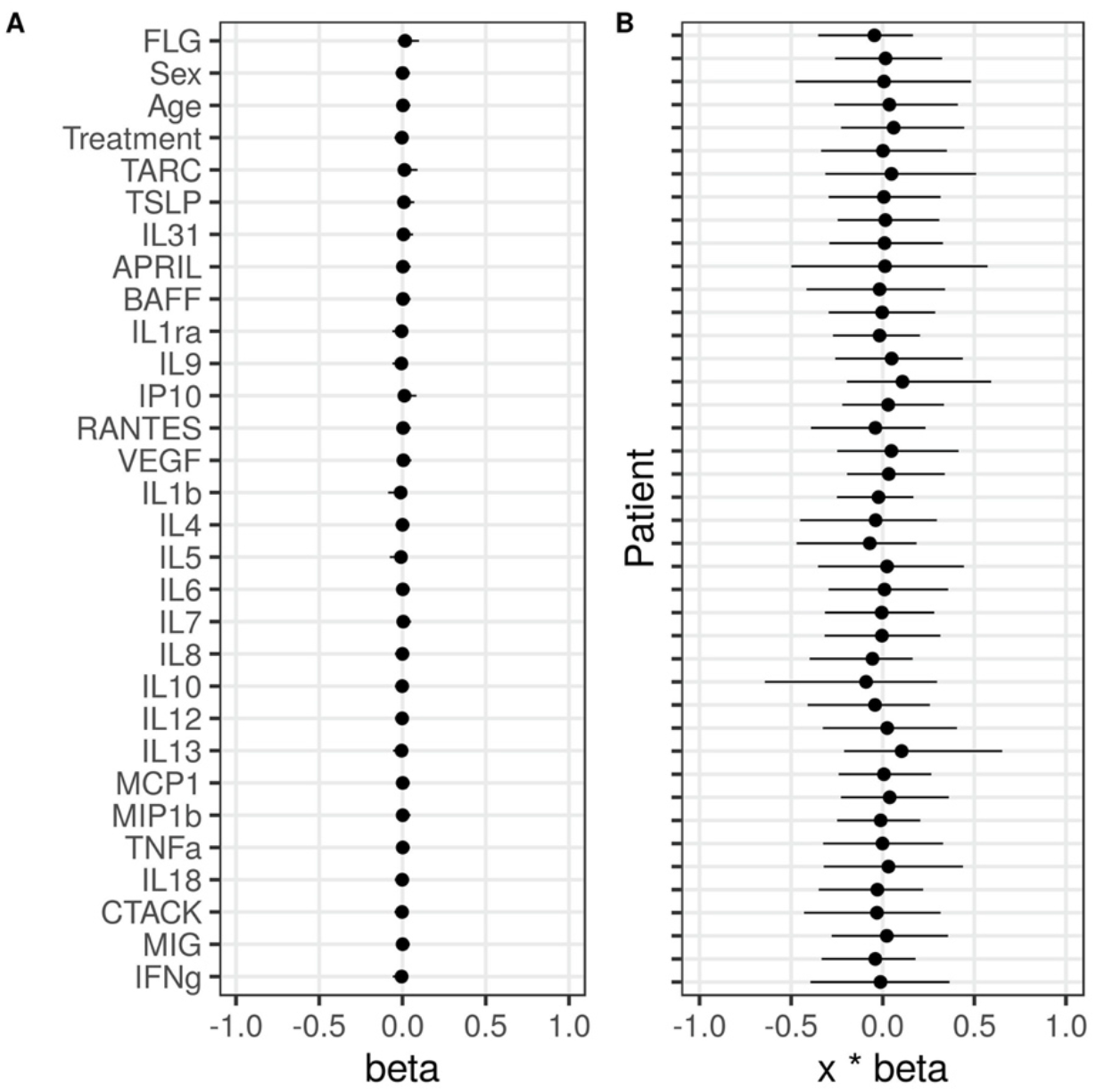
Effects of covariates in our model’s predictions of EASI (mean and 90% credible intervals). **A:** Estimates of the coefficients for the 26 serum biomarkers and FLG, sex, age and the treatment applied. A change of one standard deviation in the covariate corresponds to a change of 1.0 in EASI score. **B:** Total contribution of all covariates (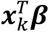) to EASI prediction for each patient.

## DISCUSSION

We developed a Bayesian state-space model with covariates that can predict AD severity scores (EASI, SCORAD, oSCORAD and POEM) two-weeks ahead in the future at the individual level. The model describes the dynamics of the latent severity and the measurement process of the severity scores (Fig. 1). The model was trained on the data from 42 adult AD patients who received systemic therapy in a previously published clinical study [13] [14] (Fig. 2). Our model outperformed reference models for time-series forecasting (Fig. 3) and revealed that biomarkers’ concentrations measured before the start of the therapy did not carry information for prediction of future AD severity scores, the primary outcomes when we evaluate the efficacy of the therapy (Fig. 4).

These results provide some insights into the sought-after roles of biomarkers in prediction of therapeutic responses, while validation on different cohorts of patients is still required. We showed that the prediction error of future scores is primarily due to errors in the score measurement process, suggesting that the effect of (any combinations of) biomarkers on a prediction of severity scores, if any, is likely to be small or too subtle to be captured by our model. Further investigation of the effect of biomarkers therefore requires the data from a larger cohort.

However, it is unclear how much new information we can expect to obtain by the inclusion of more biomarkers, as the biomarkers included in this study have been claimed to be most related to AD [4] and biomarkers usually demonstrate high multicollinearity between them. Instead, frequent measurements of biomarkers are required to evaluate their consistent effects to harness patient stratification [23]. The biomarkers’ concentrations measured at a single timepoint are likely to be noisy and may not capture the dynamic heterogeneity of complex disease such as AD. Whether the benefit of potentially more accurate predictions with biomarkers outweighs the cost of collecting data for such models remains as an open question.

A key feature of our proposed model is the quantification of uncertainties in the model parameters by full Bayesian inference, which is especially suitable when dealing with small datasets. Bayesian inference provides a flexible modelling framework to develop bespoke models, for example, by using a regularised horseshoe prior to introduce sparsity in the regression parameters while avoiding a conservative feature selection. We also modelled uncertainties in the measurements, offering a flexible way to deal with missing values or observations at irregular intervals. Missing values were treated as an absence of the measurement process in a semi-supervised learning setting. Similar models could be developed to study the dynamic evolution of AD severity scores. For example, it will be interesting to fit our model to data with more frequent measurements to investigate the short-term dynamics of AD severity scores. We could also investigate the effects of other covariates, such as air pollutants and environmental factors, that are thought to be associated with AD development and aggravation [25].

Modelling the measurement process allowed us to estimate the minimum detectable change (MDC) for the AD severity scores. The estimated MDCs suggested that it may be easier to predict objective scores such as EASI and (o)SCORAD than subjective scores such as POEM, as the MDC for EASI and (o)SCORAD was estimated to be approximately 11% of their respective range, while that for POEM was 27% of its range. Our MDC estimates are larger than already published estimates of the minimal clinically important difference (MCID) for EASI, (o)SCORAD and POEM [24] (Table S2), indicating that the changes in an outcome that a patient may identify as important are not always detectable. Further research is needed to elucidate where this difference comes from and how we ensure that clinically important changes cannot be attributed to measurement errors.

Our results suggest that a historical record of severity scores, rather than biomarkers measurements, provides rich and insightful dynamical information for prediction of therapeutic responses. While biomarkers could be used as substitutes to AD severity scores, measuring them may be costly, slow and inconvenient for routine cares. Using tools for auto-evaluation of AD severity scores from camera images, such as EczemaNet [26], may be another alternative to reduce systematic errors due to inter- and intra-rater variability in score measurement.

## Data Availability

All the codes are available at https://github.com/ghurault/ssm-eczema-biomarkers

## DATA AVAILABILITY

All the codes are available at https://github.com/ghurault/ssm-eczema-biomarkers

