## Supporting Information for "Can serum biomarkers predict the outcome of systemic therapy for atopic dermatitis?"

### A. Priors for Bayesian state-space model (SSM)

We chose the priors for our SSM to be weakly informative. Weakly informative priors are priors designed to rule out unreasonable parameter values (e.g. noise parameters outside the range of the scores) without excluding any values that could make sense. The influence of weakly informative priors is expected to disappear with enough data. We confirmed that our priors were reasonable by conducting prior predictive checks and that our results were not sensitive to the choice of realistic priors.

Instead of defining priors for $\sigma_{\mathrm{m}}$ and $\sigma_{\mathrm{l}}$, we parametrised the model with $\sigma_t = \sqrt{\sigma_{\mathrm{m}}^2 + \sigma_{\mathrm{l}}^2}$, the standard deviation for two-weeks ahead prediction and $\rho^2 = \frac{\sigma_{\mathrm{m}}^2}{\sigma_{\mathrm{t}}^2}$, the ratio of the measurement variance on the total variance. $\rho^2$ can be interpreted similarly to an R-squared, the proportion of the explained variance (the variance of the measurements) in the total variance. The priors for $\sigma_{\mathrm{t}}$ and $\rho^2$ are given by

- $\frac{\sigma_{\mathrm{t}}}{M} \sim \log N(-\log 20, (0.5 \log 5)^2)$, a lognormal prior with a 95% confidence interval of $[0.01M, 0.25M]$, and
- $\rho^2 \sim Beta(4, 2)$, a Beta distribution to reflect our expectation that future severity scores are predictable ($\sigma_{\mathrm{l}} < \sigma_{\mathrm{m}}$).

We assumed a hierarchical prior for the autocorrelation parameter, $\alpha_k \sim \mathrm{Beta}(\mu_\alpha \phi_\alpha, (1-\mu_\alpha)\phi_\alpha)$, where $\mu_\alpha$ is the population mean of the Beta distribution and $\phi_\alpha$ is the pseudo population sample size of the Beta distribution. The priors for $\mu_\alpha$ and $\phi_\alpha$ are given by

- $\mu_\alpha \sim \mathrm{Beta}(2, 2)$, a Beta distribution that slightly favours values around 0.5 as opposed to 0 or 1, and
- $\phi_\alpha \sim \log N(\log 10, (0.5 \log 10)^2)$, a log-normal prior with a 95% confidence interval being approximately $[1, 100]$, allowing a wide variety of distributions for $\alpha_k$ from well spread to concentrated.

We defined the prior for the intercept, $b_k$, by introducing the expected value of the autoregressive process, $S_k^\infty$, such that $b_k = (1-\alpha_k)S_k^\infty$. We assumed a Gaussian hierarchical prior on $S_k^\infty \sim N(\mu_\infty, \sigma_\infty^2)$, where $\mu_\infty$ is the population mean of $S_k^\infty$ and $\sigma_\infty$ is the population standard deviation of $S_k^\infty$. The priors for $\mu_\infty$ and $\sigma_\infty$ are given by

- $\frac{\mu_\infty}{M} \sim N(0.5, 0.25^2)$, a Gaussian distribution that covers the range $[0, M]$ of the score, and
- $\frac{\sigma_\infty}{M} \sim N^+(0, 0.125^2)$, a half-Gaussian distribution to reflect an assumption that $\sigma_\infty$ is at most $0.25\ M$, resulting in the width of the distribution for $S_k^\infty$ to be at most $M$.

We assumed a regularised horseshoe prior for the coefficients, $\beta_i$ ($i$=1, …, 30 = $D$), defined by

- $\beta_i \sim N(0, \tilde{\lambda}_i^2 \tau^2)$, where $\tilde{\lambda}_i = \frac{c^2 \lambda_i^2}{c^2 + \tau^2 \lambda_i^2}$ is the local shrinkage parameter, $\tau$ is the global shrinkage parameter and $c$ is the scale of the signal,
- $\lambda_i \sim C^+(0, 1)$, where $C^+$ denotes a half-Cauchy distribution,
- $\tau \sim C^+\left(0, \frac{p_0}{D-p_0}\frac{\sigma}{\sqrt{N}}\right)$, where $p_0 = 5$ is the expected number of covariates with non-zero coefficients, $D = 30$ is the number of covariates, $N = 42$ is the number of patients, and $\sigma = \sigma_{\mathrm{l}}$ is the standard deviation of the residuals, and
- $c^2 \sim \text{Inv-}\chi^2(\nu, \sigma_\chi^2)$, a scaled-inverse chi-squared prior, where we assume the degree of freedom, $\nu = 5$, and the scale, $\sigma_\chi = 1$. This prior corresponds to $c$ following a Student-t slab with $\nu$ degrees of freedom and scale $\sigma_\chi$, if $\beta_i$ is far from 0. This prior reflects an assumption that the order of magnitude of non-zero coefficients is around 1 but could be higher.

To avoid the situation where most of the mass of the non-truncated distribution for the latent score, $\hat{S}_k(t)$, are outside of the range $[0, M]$, which can cause computational problems, we implemented a soft-uniform prior on $\hat{S}_k(t)$. The soft-uniform prior is defined by the probability density function, $f(x) = \frac{logit^{-1}(x+0.01M) - logit^{-1}(x-1.01M)}{1.02M}$, resulting in an approximately constant density between 0 and $M$ (i.e. not prioritising any values in this range) with a slow convergence to 0 (i.e. penalising values) outside this range.

### B. Reference models

We implemented four reference models, a uniform forecast model and three models of increasing complexity leading to our state-space model. The models were implemented in a Bayesian setting and provided probabilistic predictions for a fair comparison. These models are more advanced than standard off-the-shelf implementation as missing values are treated as parameters to be inferred in a semi-supervised setting.

- The uniform forecast model is described by $S_k(t) \sim U(0, M)$, where each outcome is assigned the same probability density.
- The random walk model provides a flat forecast, $S_k(t+1) \sim N(S_k(t), \sigma^2)$, centred on the last observation with the uncertainty quantified by a variance, $\sigma^2$. The prior for $\sigma$ is the same as that for $\sigma_\mathrm{t}$ in our SSM.
- The autoregressive model is an extension of the random walk model and is described by $S_k(t+1) \sim N(\alpha\, S_k(t) + b, \sigma^2)$, with a fixed autocorrelation, $\alpha$, and an intercept, $b = (1-\alpha)S^\infty$. We assumed a uniform prior for $\alpha \sim U(0,1)$. The prior for $S^\infty$ is the same as that for $\mu_\infty$ in our SSM.
- The mixed effect autoregressive model extends the autoregressive model and is described by $S_k(t+1) \sim N(\alpha_k\, S_k(t) + b_k, \sigma^2)$, with a patient-dependent autocorrelation, $\alpha_k$, and a patient-dependent intercept, $b_k = (1-\alpha_k)S_k^\infty$. The priors for $\alpha_k$ and $b_k$ are the same as those in our SSM.

*Table S1: Posterior summary statistics of the population-level parameters for the model predicting EASI without covariates.*

| Parameter | Interpretation | $\hat{R}$ | $N_{\mathrm{eff}}$ | Mean | SD | 2.5% | 50% | 97.5% | PS* |
|---|---|---|---|---|---|---|---|---|---|
| $\sigma_t$ | Square root of the total variance | 1.000 | 1760 | 4.499 | 0.283 | 3.976 | 4.488 | 5.094 | 0.988 |
| $\rho^2$ | Proportion of the $\sigma_\mathrm{m}^2$ in the total variance | 1.002 | 1055 | 0.947 | 0.037 | 0.853 | 0.955 | 0.993 | 0.955 |
| $\sigma_\mathrm{l}$ | Standard deviation of the latent dynamic | 1.003 | 1089 | 0.968 | 0.340 | 0.371 | 0.946 | 1.671 | 0.939 |
| $\sigma_\mathrm{m}$ | Standard deviation of the measurement process | 1.001 | 1453 | 4.379 | 0.310 | 3.794 | 4.372 | 5.013 | 0.982 |
| MDC | Minimum Detectable Change for a 95% confidence level | 1.001 | 1453 | 8.582 | 0.607 | 7.436 | 8.568 | 9.826 | 0.982 |
| $\mu_\alpha$ | Population mean of the autocorrelation parameter $\alpha$ | 1.002 | 1572 | 0.688 | 0.042 | 0.600 | 0.691 | 0.765 | 0.961 |
| $\phi_\alpha$ | Population "pseudo sample size" of the autocorrelation $\alpha$ | 1.004 | 1174 | 3.664 | 1.094 | 2.026 | 3.504 | 6.187 | 0.999 |
| $\mu_\infty$ | Population mean of the autoregressive process | 1.002 | 1597 | 3.574 | 0.588 | 2.446 | 3.561 | 4.789 | 0.997 |
| $\sigma_\infty$ | Population standard deviation of the autoregressive process | 1.002 | 2017 | 0.507 | 0.405 | 0.017 | 0.421 | 1.489 | 0.986 |

* The Posterior Shrinkage (PS) of the parameter $\theta$ is defined as $1 - \frac{\mathrm{Var}(\theta_{\mathrm{post}})}{\mathrm{Var}(\theta_{\mathrm{prior}})}$. PS near 0 indicates that the data provides little information beyond the prior and PS near 1 indicates that the data is much more informative than the prior.

*Table S2: MCID reported in* [24] *and MDC estimated in this study*

| | EASI | SCORAD | oSCORAD | POEM |
|---|---|---|---|---|
| **MCID [24]** | 6.6 | 8.7 | 8.2 | 3.4 |
| **MDC (mean and 90% credible interval)** | 8.6<br>[7.6, 9.6] | 11.4<br>[9.1, 13.5] | 9.1<br>[7.4, 10.7] | 7.7<br>[6.7, 8.9] |

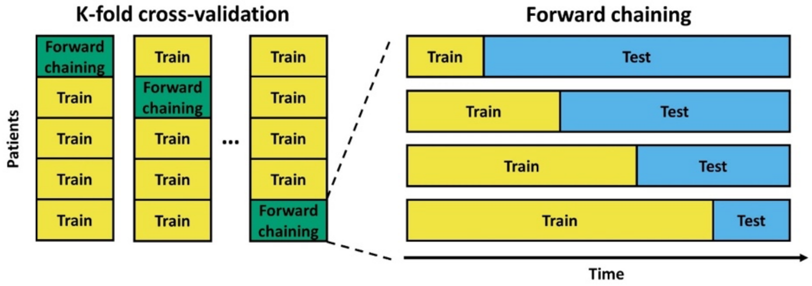


*Figure S1: K-fold cross-validation ($K = 5$) in a forward chaining setting.*

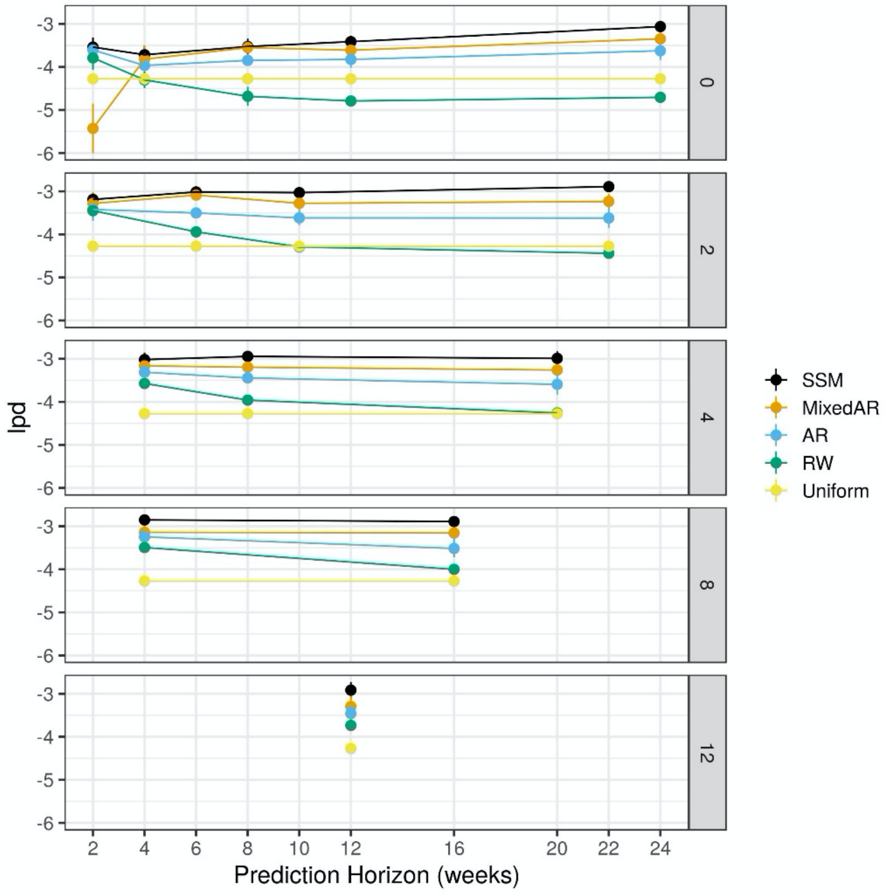


*Figure S2: The values of lpd (mean ± SE; the higher the better) as a function of the prediction horizon for various training weeks (panels) and models (colours) predicting EASI.*

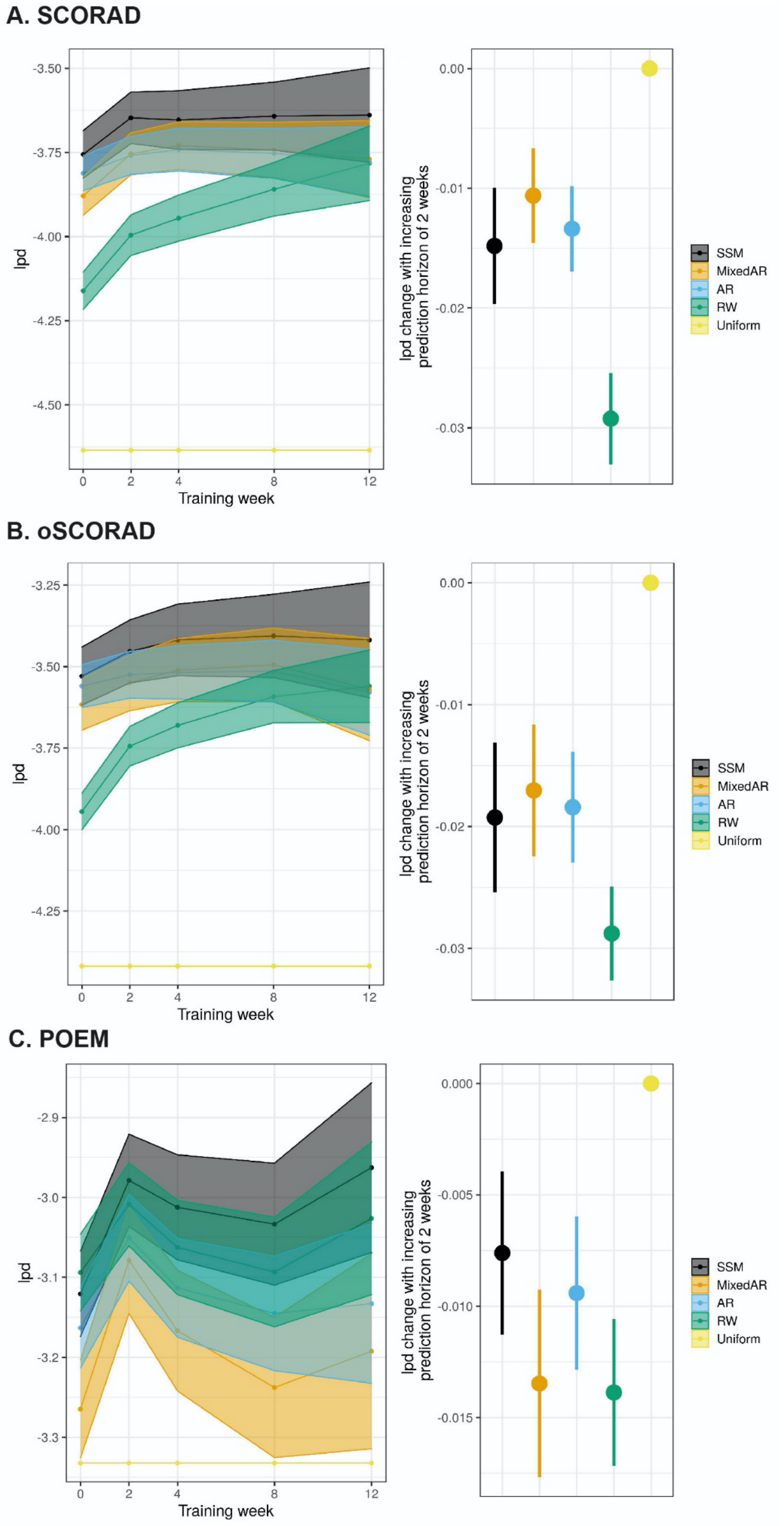


*Figure S3:* Predictive performance of our model (SSM) and reference models (MixedAR, AR, RW and Uniform) for oSCORAD (**A**), SCORAD (**B**) and POEM (**C**). The performance was evaluated by lpd (higher the better). Left: Learning curves (mean ± SE) for two-weeks ahead prediction after adjusting for different prediction horizons. Right: Change in lpd as the prediction horizon is increased by two weeks.
